# Transcriptome-Wide Alternative Splicing Analysis Implicates Complex Events in Bipolar Disorder

**DOI:** 10.64898/2026.04.19.26351209

**Authors:** Miriam Martínez-Jiménez, Inés García-Ortiz, Diego Romero-Miguel, Tomas Kavanagh, Lee L Marshall, Rosa Ana Bello Sousa, Sergio Sanchez Alonso, Raquel Álvarez García, Sergio Benavente López, Ezequiel Di Stasio, Peter R Schofield, Enrique Baca-García, Philip B Mitchell, Antony A Cooper, Janice M Fullerton, Claudio Toma

## Abstract

Alternative-splicing events (ASE) increase transcriptomic variability and play key roles in biological functions. The contribution of ASE to bipolar disorder (BD) remains largely unexplored. We performed a Transcriptome-Wide Alternative-Splicing Analysis (TWASA) to identify ASEs and genes potentially involved in BD.

The study comprised 635 individuals: a discovery sample (DS) of 31 individuals from eight multiplex BD families (16 BD cases; 15 unaffected relatives), and a replication sample (RS) of 604 subjects (372 BD cases; 232 controls). Sequencing was conducted on RNA from lymphoblastoid cell lines (DS) and whole blood (RS). TWASA was performed using VAST-TOOLS (VT), rMATS (RM), and MAJIQ/MOCCASIN (MCC). Gene-set association analyses of genes containing ASEs were performed across six psychiatric disorders. Novel ASE (nASE) were investigated in the DS using FRASER.

Limited gene overlap was observed across TWASA tools. MCC identified 2,031 complex ASEs involving 1,508 genes, showing the strongest genetic association with BD across psychiatric phenotypes. Prioritization of MCC-identified ASE genes yielded 441 candidates, including *DOCK2* as top candidate from the DS. Replication was obtained for 98 genes, five with an identical ASE, and four (*RBM26*, *QKI*, *ANKRD36*, and *TATDN2*) showing a concordant percentage-spliced-in direction with the DS. Finally, 578 nASE were identified in the DS, with no evidence of familial segregation or differences in ASE types. This first TWASA in BD reveals tool-specific variability, complex ASE for genes specifically associated with BD, and novel candidate genes for BD. Alternative transcript isoform abundance may represent a mechanism contributing to BD pathophysiology.

## INTRODUCTION

Bipolar disorder (BD) is a chronic psychiatric condition characterized by recurring manic and depressive episodes [1], affecting approximately 1–2% of the global population [2,3]. Genetic factors play a substantial role in disease aetiology [4], as observed by strong familial aggregation and heritability estimates of up to 80% [5]. Genome-wide association studies (GWAS) and next-generation DNA sequencing studies have considerably advanced the identification of genetic susceptibility factors for BD. The most recent GWAS meta-analysis conducted by the Psychiatric Genomics Consortium (PGC) identified 298 genome-wide significant loci [6], supporting the highly polygenic architecture of the disorder. Additionally, case-control sequencing initiatives, although more limited in scale than GWAS, have begun to implicate candidate genes harbouring rare variants [7]. Furthermore, sequencing studies of large extended BD families suggest a complex pattern of inheritance, with rare variants likely to exert modest penetrance effects compared to previous estimates [8]. Notably, recent evidence shows an increased polygenic burden of common risk alleles, together with contributions from *de novo* variants, even within multiplex families [9,10]. These findings support a model in which BD likely results from the cumulative effects of hundreds of risk alleles across different types of genomic variation, including inherited and *de novo* variants, implicated in both sporadic cases and multiplex families [7,9,10].

Recently, an integrative fine-mapping strategy applied to a BD GWAS successfully prioritized 17 putative causal single-nucleotide polymorphisms (SNPs) and their corresponding genes from significant GWAS loci [11]. However, only a small proportion of these SNPs identified through GWAS are predicted to directly affect protein-coding sequences, while most are located in non-coding regions, and may therefore act on gene expression or regulation.

Alternative-splicing represents a key mechanism of gene regulation, allowing a single gene to generate multiple transcripts, and consequently, protein isoforms with distinct or even divergent biological functions [12]. Alternative-splicing events (ASE) arise through the selective inclusion or exclusion of exons from a pre-mRNA transcript, generating diverse mRNA isoforms and substantially expanding the proteomic and functional complexity of the genome [13]. In addition to constitutive splicing, ASEs are commonly classified into five main categories: cassette alternative exon, alternative 5’ splice site, alternative 3’ splice site, mutually exclusive exons, and intron retention [14]. However, several of these basic categories may co-occur within the same transcript region, giving rise to complex ASEs that involve combinations of basic ASE categories. These complex events further increase transcriptomic diversity and generate highly intricate isoforms [15]. Notably, isoforms generated through both basic and complex ASEs can coexist, resulting in the presence of multiple distinct transcripts from the same gene within the same cell or tissue [16,17], typically at varying levels of abundance.

Brain-expressed genes are particularly enriched for ASEs, indicating that increasingly complex splicing programs evolved in parallel with vertebrate brain complexity [18,19]. In BD, one of the clearest example of ASE is represented by *ANK3*, one of the most replicated genes in BD, identified by both GWAS and sequencing studies [20,21]. Notably, a genetic variant leading to exon skipping in a specific *ANK3* isoform has recently been suggested to contribute to BD risk [22].

While several transcriptomic studies in BD have examined differences in gene expression between cases and controls [23], or affected and unaffected relatives [24], comprehensive studies of global splicing dysregulation using transcriptome-wide alternative splicing analysis (TWASA) remain extremely limited. Most TWASA in psychiatry have focused on autism spectrum disorder (ASD), schizophrenia (SCZ), major depressive disorder (MDD), or alcohol and substance use disorder, typically using a sample size smaller than 50 patients [25–37]. Nonetheless, additional large-scale TWASAs have been performed in MDD and alcohol use disorder [38–40]. One of the largest studies in this field investigated basic ASE comparing controls (N=936) with ASD (N=51), SCZ (N=559), and BD (N=222) [40]. This study reported higher rates of exon skipping in BD compared with ASD and SCZ, as well as disorder-specific splicing alteration. However, its interpretation is limited by the use of a single computational method for TWASA, and by heterogeneity arising from the inclusion of multiple brain cohorts and sequencing platforms. Similarly, an analysis of intron retention in iPSC-derived neuronal precursor cells from nine subjects, provided a preliminary insight into alternative splicing under mood stabiliser treatment [41].

Both the development of RNA sequencing (RNAseq) technology and improvement of computational tools have allowed better detection and quantification of ASEs. Such tools rely on distinct methodological frameworks and statistical models, which can lead to variability in results [42–44]. While most tools are designed to detect basic ASE, few are capable of capturing complex ASEs. These methods quantify the relative inclusion of annotated exons or introns, expressed as the percentage-spliced-in (ψ or PSI), and compare differences between cases and controls, denoted as ΔPSI (dPSI). Beyond basic and complex ASE, transcript variability also encompasses novel ASEs (nASEs), which correspond to splicing events that are novel and unannotated. This category is of particular interest, as nASEs may arise from deep functional intronic changes or disruption of canonical splice sites, potentially generating pathogenic transcript isoforms. Despite the relevance of nASEs, computational tools capable of reconstructing novel transcript structures remain limited, and their application to psychiatric disorders has been largely unexplored.

To address these gaps, the present study aimed to compressively investigate global splicing dysregulation in BD. We analysed basic ASEs, complex ASEs, and nASEs using multiple computational pipelines. A discovery analysis was conducted in a family-based cohort, followed by replication of candidate ASE genes in an independent case-control cohort, enabling identification of candidate genes for splicing alterations in BD.

## METHODS AND MATERIALS

### Study cohorts and clinical assessments

The TWASA was conducted in the discovery sample (DS) comprising eight multiplex BD families, each with at least three affected individuals across two generations [24]. Family members included individuals diagnosed with BD type-I (BD-I), BD type-II (BD-II), schizoaffective disorder-manic type (SZMA) as well as unaffected relatives. Participants underwent clinical assessment via the “Family Interview for Genetic Studies” (FIGS) [45] and the “Diagnostic Interview for Genetic Studies” (DIGS) [46]. The replication sample (RS) consisted of patients with BD-I, BD-II, and SZMA patients (N=372) and controls (N=232) from the Madrid Manic Group (MadManic) cohort [47]. Additional clinical information is provided in the Supplementary material.

### RNA sequencing and Preprocessing

For the DS, total mRNA was extracted from lymphoblastoid cell lines of 31 individuals (16 BD cases; 15 unaffected relatives). Cell culture, RNA extraction, and RNAseq were performed as previously described [24]. For the RS, total RNA was extracted from whole blood, and globin depleted. RNAseq was conducted using the NovaSeq X Plus platform (Illumina). Raw reads were quality checked using FastQC v0.11.9 [48], and then trimmed using Trimmomatic v0.39 [49]. Post-trimming quality checks were performed using default parameters. Reads were aligned to the GRCh38.p13 reference genome using Hisat2 v2.1.0 [50], and resulting .bam files were sorted and indexed using SAMtools v1.9 with default arguments [51]. Additional details on RNA processing and sequencing are provided in the Supplementary material.

### Alternative Splicing computational tools

The TWASA was performed by comparing BD cases with their unaffected relatives (DS), using three different computational tools: VAST-TOOLS v2.5.1 (VT) [52], rMATS v4.1.2 (RM) [53], and a more recently developed combined pipeline integrating MAJIQ and MOCCASIN v2.4 (MCC) [54,55]. While VT and RM detect only basic ASEs, MCC is capable of identifying both basic and complex ASEs.

For replication purposes, sporadic BD cases and unrelated controls (RS) were analysed with MCC only. The overall workflows are summarized in **Fig. 1**.

**Figure 1:**
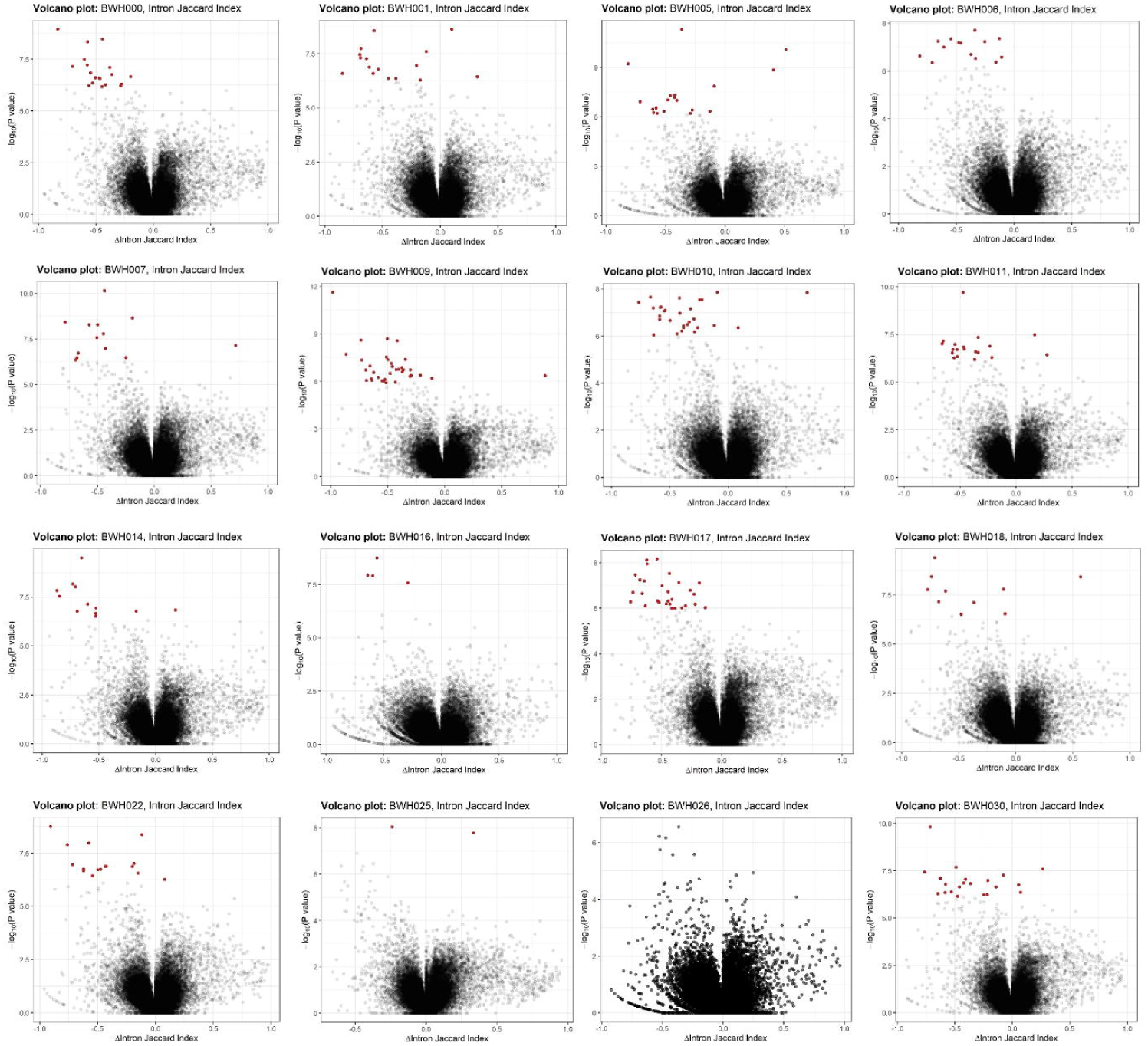
Overview of the TWASA pipelines used in this study. Alternative-splicing events (ASE) were assessed between groups using three computational approaches: VAST-TOOLS v2.5.1 (VT) in blue, rMATS v4.1.2 (RM) in red, and the combined MAJIQ and MOCCASIN v2.4 pipeline (MCC) in green. *Novel* ASEs (nASEs) were identified at individual level using FRASER in purple. For all splicing analyses, FASTQ files served as input and were processed through the respective pipelines to generate TSV files containing raw ASE results.

VT was executed sequentially using the *align, combine*, and *tidy* modules. Additional methodological details are provided in supplementary material. For RM, the STAR indexes were generated from FASTQ files using STAR v2.7.0d [56]. RM was executed with default parameters, and the *Homo_sapiens.GRCh38.107.chr.gtf* annotation file from Ensembl.

The MCC pipeline was performed first using the *Majiq build* module with sorted .bam and .bai files, together with the *Homo_sapiens.GRCh38.107.gff3* annotation file, to detect local splicing variation (LSV) candidates. This step also generates the splice graph (.sql file) and the indexes (.majiq files) required for the subsequent MAJIQ analyses, which was run with the *--simplify 0.01* flag. The build files were then corrected using MOCCASIN [55], via *moccasin.py* script, which takes the .majiq files and a model matrix containing confounding covariates to generate *.scf_adjusted.majiq* files. In the DS, gender, age, and familyID were included as covariates; while for the RS, gender and age were used. No intercept column was included in either model matrix.

For ASE quantification in MAJIQ the *heterogen* module was used for group comparisons. Input files (.majiq or the .scf_adjusted.majiq) were analyzed with the *--min-experiments 2* flag and *P-values* calculated using the *--stats WILCOXON* option. Final ASE results were generated using the *voila tsv* with the *--changing-between-group-dpsi 0.05* flag, and visualized via the *voila view* as HTML files.

From TSV output of all three tools, ASEs were filtered using |dPSI| > 5% and *P-value* < 0.05. An additional filter for higher coverage, defined as ≥ 30 reads per individual for the significant alternative splicing sub-event, was applied to the MCC TSV output. The same settings were used in the RS for MCC pipeline. To reduce false positive in the replication phase only, ASEs identified in ≥90% of subjects in both BD or control groups were considered, together with |dPSI| > 5% and *P-value* < 0.05.

The MAJIQlopedia database was consulted to assess whether ASEs occurred in other tissues [57]. The ExPASy tool [58] was used to predict the amino acid sequences of proteins resulting from ASEs of interest. Phosphorylation of threonine residues was predicted using NetPhos 3.1 [59]. Additional tool-specific details are provided in the Supplementary material.

### Novel alternative splicing events (nASE) analysis

FRASER v2.0 [60], implemented within the DROP v1.4 workflow [61,62], was used to identify nASEs as summarized in **Fig. 1**. The following parameters were used in the DROP under *aberrantSplicing* section: i) run: true; ii) groups: together; and iii) a minimum read threshold: ≥30 reads in at least one sample. FRASER incorporates a denoising autoencoder to automatically account for confounders factors. The default principal component analysis (PCA)-based option was selected for confounder correction. Comparison of nASE categories between affected and unaffected family members was performed via Wilcoxon test.

### Gene-based and gene-set association analysis

MAGMA v1.09 was used to perform gene-based association (GBA) and gene-set association analyses (GSA) [63]. For GBA, a window of 5kb downstream and upstream for each gene was considered, and associations were tested using a multi-SNP wise model. GSA was performed using gene lists derived from RM, VT and MCC following ASE filtering, and employed gene-based *P-values* calculated from coding genes only, for GWAS summary statistics of six psychiatric conditions (**Table S1**).

### Functional enrichment analysis and in-silico validation via MAGMA GSA

An over-representation analysis (ORA) was performed on the MCC-derived gene list to assess enrichment in biological functions. The *enricher* function from the *clusterProfiler* R package (v. 3.16) [64] was applied using categories from the Molecular Signature Databases (MSigDB, msigdbr R package, v. 7.5), including the following curated datasets: i) Gene Ontology (GO), encompassing Molecular Function, Biological Processes, and Cellular Component; ii) Human Phenotype Ontology (HPO); iii) Reactome; and iv) Kyoto Encyclopedia of Genes and Genomes (KEGG) [24].

Enriched categories were subsequently evaluated for their association with BD and SCZ using GSA based on GWAS summary statistics of these conditions (**Table S1**). To identify and visualize categories shared between BD and SCZ, a bubble plot was generated using an in-house implementation of the *GOBubble* function from *GOplot* R package (v. 1.0.2) [65].

## RESULTS

### Comprehensive alternative splicing analyses through multiple computational tools

TWASA was performed in the DS using three independent tools, namely VT, RM, and MCC, to identify ASEs and assess consistency across analytical approaches. VT identified 402 genes and 422 ASEs (**Table S2**). Although none of these ASEs survived multiple-testing correction, the most strongly associated genes included *HTT* (*P*=1.39E-04), *ZFYVE16* (*P*=1.54E-04), and *ABCD4* (*P*=5.76E-04). RM detected 297 genes and 354 ASEs **(Table S3**), of which 32 ASEs in 22 genes remained significant after FDR correction. Notably, significant ASEs were identified in *TTLL3* (*P*=4.3E-12; *Padj*=6.05E-07), *NPM2* (*P*=6E-10; *Padj*=4.24E-05), and *RPS6KL1* (*P*=2.5E-09; *Padj*=1.99E-05). MCC identified a substantially larger set of 1,508 genes and 2,031 ASEs **(Table S4**), with top-ranked ASEs observed in *MICAL1* (*P*=1.78E-05), *GOLGA3* (*P*=2.29E-05), and *LRCH3* (*P*=7.34E-05).

Overlap of genes and ASEs across the three TWASA approaches is illustrated in **Fig. 2A**. Notably, no genes or ASEs were consistently identified across all three tools (**Table S5**). At the gene level, VT and MCC showed the greatest overlap, with 65 genes in common. However, only four of them corresponded to identical ASEs (**Table S5**). Twenty-one genes were shared between VT and RM, with four matching the same ASE (**Table S5**). Meanwhile, 32 genes overlapped between RM and MCC, of which only one corresponded to the same ASE (**Table S5**).

**Figure 2:**
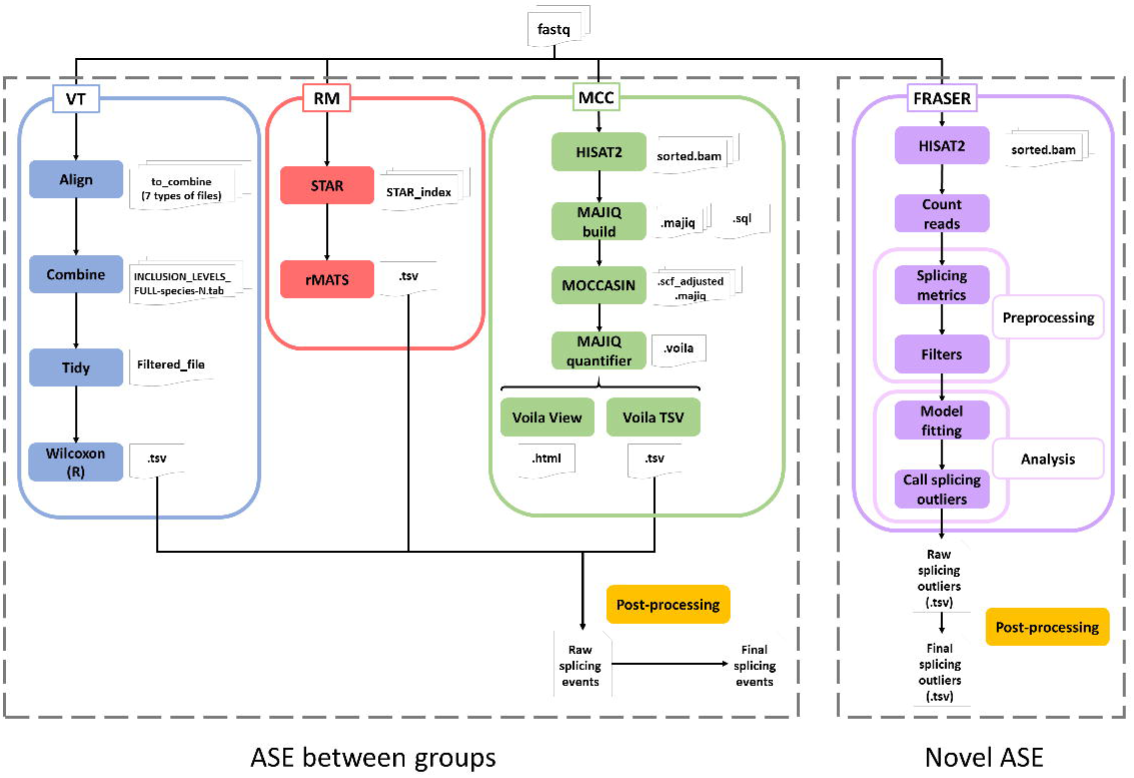
**A)** Venn diagrams illustrating TWASA concordance across the three computational tools used for ASEs detection in the DS. The left panel shows overlap at gene level, while the right panel depicts overlap at the event level. **B)** Balloon plot summarizing gene-set enrichment association results for ASE genes identified by VAST-TOOLS (VT), rMATS (RM), and MAJIQ-MOCCASIN (MCC), using a consistent threshold of deltaPSI>5 and p<0.05. Gene-set enrichment was tested using MAGMA across six psychiatric phenotypes: major depressive disorder (MDD), schizophrenia (SCZ), bipolar disorder (BD), substance use disorder (SUD), attention deficit/hyperactivity disorder (ADHD), and suicide attempt (SUI). Discovery GWAS sample size is provided in parentheses for each condition. Balloon size and colour intensity correspond to –log10(*P-value*)^3. Nominal gene-set association P-values (*P*<0.05) (Table S6) are indicated by an asterisk (*).

Next, gene-set lists derived from ASEs identified by each of the three algorithm were investigated for their combined association against six psychiatric disorders (**Fig. 2B**). Only complex ASEs identified with MCC showed statistically significant associations with BD (*P*= 0.0025), SCZ (*P*= 0.0118), and SUD (*P*= 0.0117) (**Table S6**), whereas genes containing basic ASEs identified by VT and RM exhibited no significant genetic contributions to any trait. Therefore, all subsequent analyses took into account only the ASEs identified by MCC, which comprise both basic and complex events.

### Functional enrichment and in silico validation

To investigate the functional relevance of MCC-derived ASE genes, we performed an ORA using the initial set of 1,508 genes with statistically significant ASEs. This analysis identified 1,221 enriched functional categories with Padj<0.05 (**Table S7**). To assess the robustness and disease relevance of these functional enrichments, each category was subsequently validated for genetic association enrichment using GWAS of BD and SCZ via GSA implemented in MAGMA. Out of the 1,221 categories identified, 158 were successfully associated to BD and 168 to SCZ. Notably, 51 categories showed significant enrichment in both disorders (**Fig. 3**). Among these shared categories, 22 were relevant to psychiatric conditions, including terms related to aggressive behaviour, synaptic function, and other neurobiological processes (**Fig. 3**).

**Figure 3:**
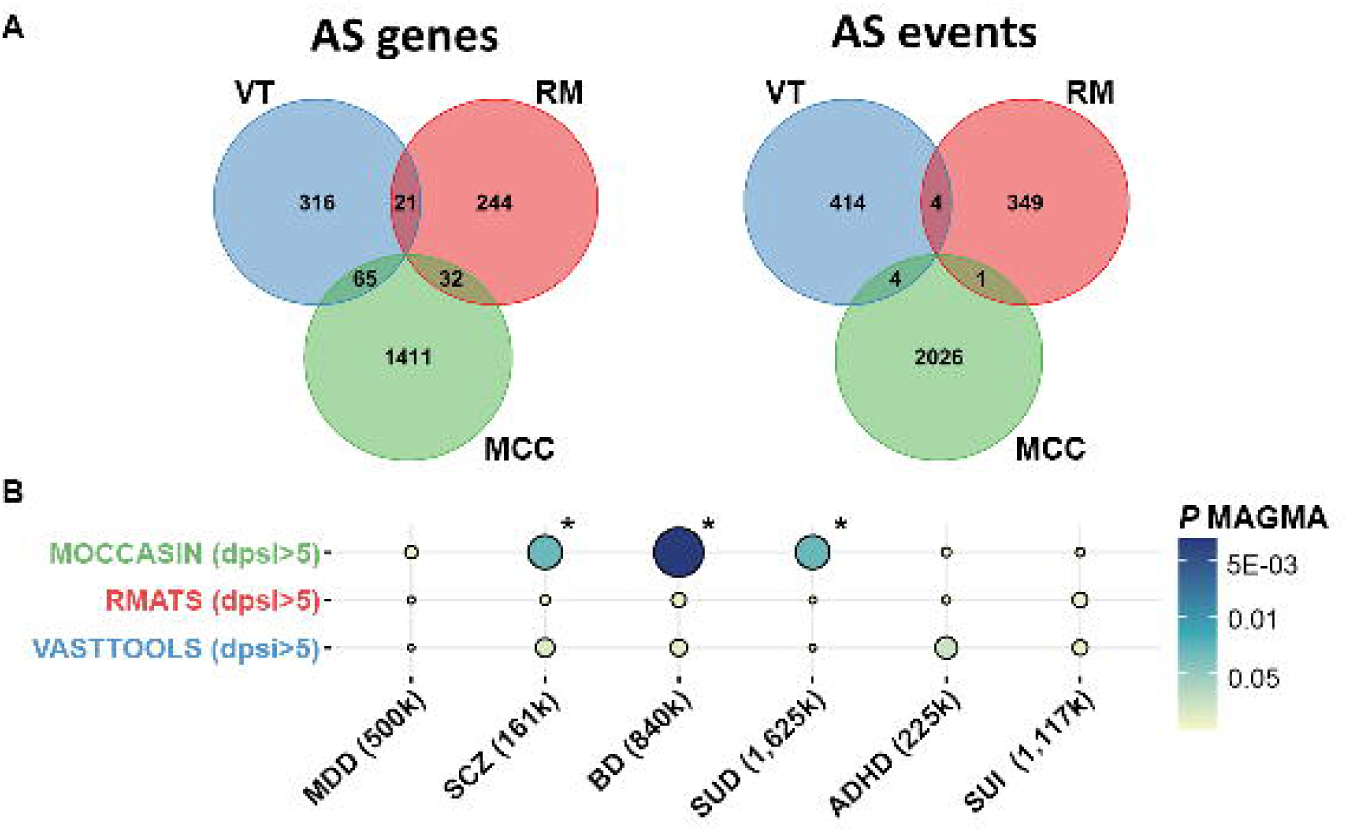
Over-representation analysis (ORA) of enriched categories derived from the ASE gene set identified by MCC. These categories were further evaluated for genetic associations via MAGMA using GWAS for BD and SCZ. Categories shared between BD and SCZ with particular relevance to psychiatry are labelled. Bubble area represents gene-set size. **A)** Shared categories showing significant associations with BD (orange bubbles). **B)** Shared categories showing significant association with SCZ (blue bubbles). For each category, the x-axis shows the MAGMA beta coefficient and y-axis represents –log10(*P* MAGMA).

### Candidate gene prioritization and replication in an independent cohort

From the 1,508 genes involving 2,031 ASEs identified by the MCC pipeline, we applied additional criteria beyond standard filtering and high-read coverage for gene prioritization: i) expression in brain tissues, defined as TPM > 0.1 (GTEx database v8 [66]); and ii) genes nominally associated (MAGMA *P*-value < 0.05) via a GBA with the largest available BD GWAS summary statistics (**Table S8**). Additionally, we calculated polygenic priority scores (PoPS) [24] for the selected putative causal genes in BD. This prioritization resulted in a final list of 596 ASEs in 441 genes (**Table S8**).

Amongst the ten most significant gene-based associations with BD, *DOCK2* showed the most significant alternative splicing sub-event, corresponding to an intron retention (*P*=0.00089). The *DOCK2* gene harbours a genome-wide significant GWAS signal, with rs10866641 as leading SNP (*P*= 2.79E-11). The retained intron (548 bp), preserving the coding reading frame, was more frequent in unaffected subjects, and was accompanied by a compensatory double exon-skipping event of two non-canonical exons, more frequent in BD cases, that corresponds to the canonical splicing of the main isoform. The retained intron isoform is predicted to lead to a premature stop codon (**Fig. S1**).

The prioritized gene list of 441 genes was subjected to replication with standard MCC settings in an independent cohort comprising 372 BD cases and 232 controls. After QC filtering, 423 genes were considered for replication, of which 98 (23.2%) showed one or more ASEs. Amongst these, five genes exhibited the same ASE identified in the DS analysis, and four showed the same splicing sub-event with concordant direction of effect across both cohorts. These four replicated ASEs were identified in *RBM26*, *QKI*, *ANKRD36*, and *TATDN2* (**Fig. 4**). Notably, *QKI* represented the most significant replicated finding (*P*=1.58E-07), identifying an increased inclusion of the seventh canonical intron in the control group. This alternative isoform (QKI-5B) appears to be expressed in brain tissues (**Fig. S2**), and is predicted to lead to a premature stop codon, while keeping the signal peptide for nuclear localization (**Fig. 5**). The QKI-5B consist in a shorter isoform of four amino acids with the last changing from alanine at position 337 to glycine (A337>G337). *In silico* prediction analyses of the protein-level consequences of this event (**Fig. S3**), indicate a reduction in phosphorylation potential at threonine 333, and a complete loss of the C-terminal threonine residue at position 339 predicted to be phosphorylated by an RSK kinase.

**Figure 4:**
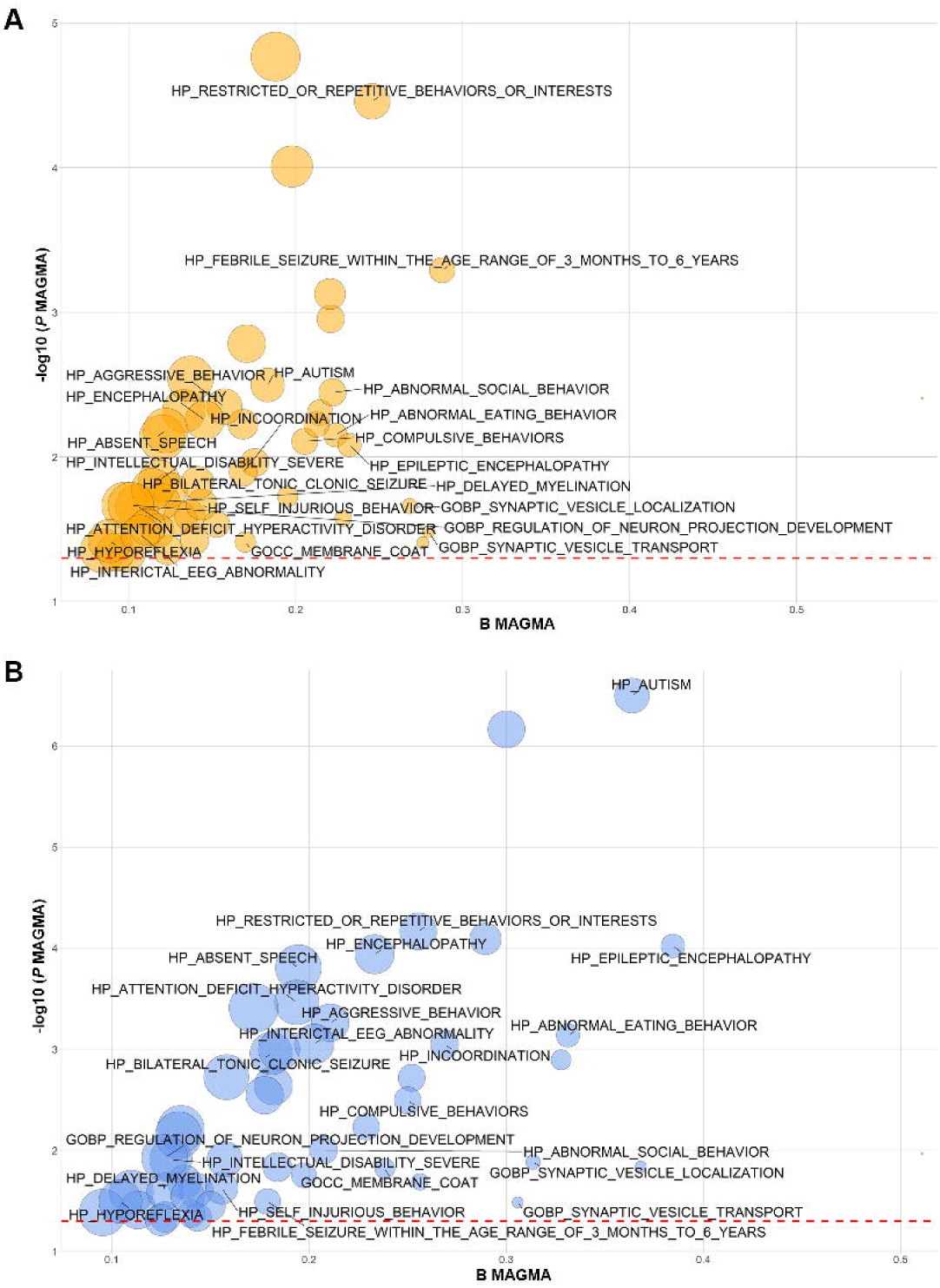
ASEs from prioritized genes identified by the MCC pipeline in the DS replicated in the RS cohort. Each panel shows above a schematic representation of the gene structure (exon/intron) with a red box highlighting the ASE location; and on the top right the splicing sub-event type in relation to exons and introns of the ASE. Below, the left side, shows violin plots from the DS (family-based) and RS (case-control) illustrating absolute dPSI values and the P-value, with colours corresponding to the sub-events shown in the top-right representation. The bottom right shows the GWAS association plot for BD for each gene. Panel A shows the replicated ASE in *RBM26* gene; B in *QKI* gene; C in *ANKRD36* gene; and D in *TATDN2* gene.

**Figure 5:**
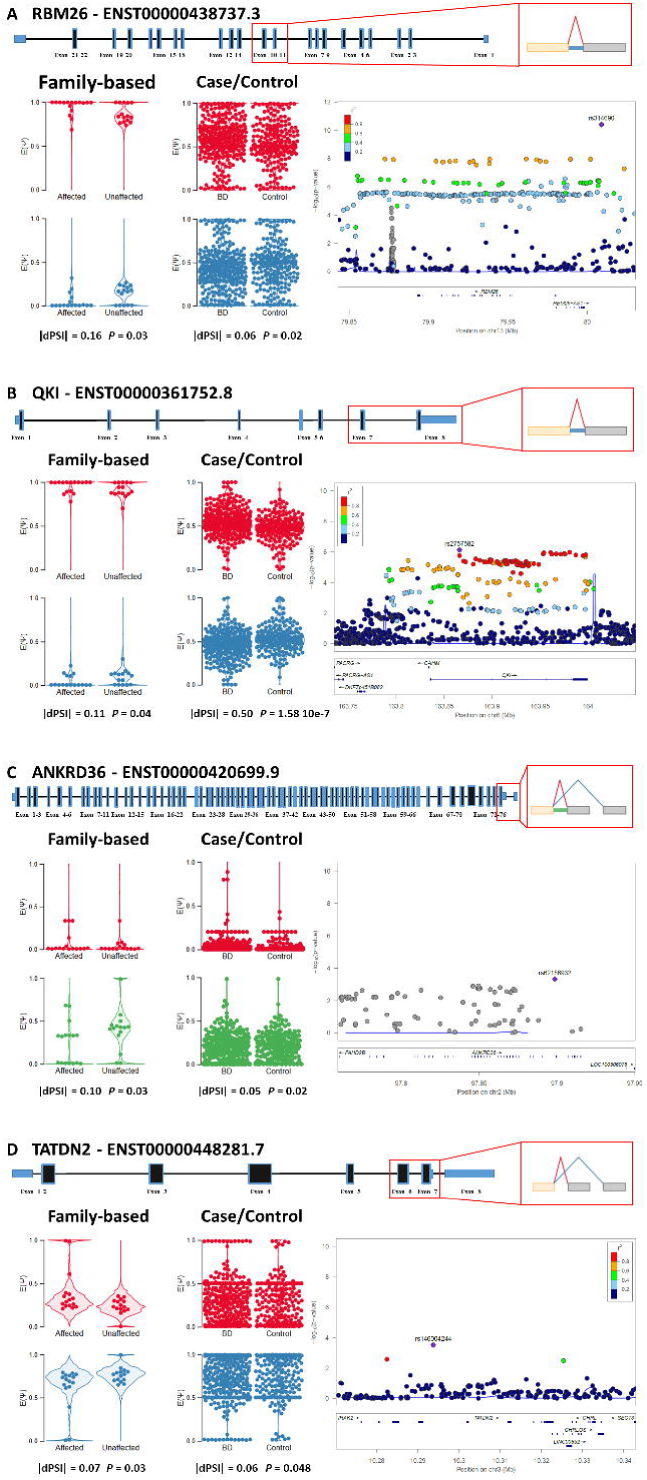
Major isoforms of the human *QKI* gene. All isoforms contain the conserved QUA1, KH, and QUA2 RNA-binding domains, while they differ in their 3′ untranslated regions (UTRs) as a result of alternative splicing and polyadenylation events. QKI-5 isoform contains a nuclear localization signal (NLS), limiting its activity to the nucleus. QKI-5B (in red) is the alternative isoform resulting from the 548 bp intron retention identified in our study. *Abbreviations:* NLS, nuclear localization signal; QKI, RNA-binding protein Quaking.

### nASE detection in the family-based cohort

To complete a holistic ASE analysis, we applied a transcriptome-wide approach to identify nASEs, which may highlight additional genes involved in BD.

On average, approximately 19 nASEs were detected per individual, ranging from zero to 36 (**Fig. S4A**). No evidence of technical inflation was observed, and the detected events showed highly significant outlier signals (**Fig. S4B**). In total, 578 significant nASEs were identified in 479 genes (**Table S10**), including 228 events in BD cases (**Fig. 6**) and 313 events in unaffected relatives (**Fig. S5**). Analysis of segregation patterns within family members revealed no clear evidence for the inheritance of family-specific nASEs. Amongst the nASEs, four were predicted to introduce frameshifts: two in BD cases (*RPL23* and *IGHM*) and two in unaffected individuals (*RPL7A* and *RPS9*) (**Table S10**). Overall, comparison of eleven nASE event types between BD cases and unaffected relatives revealed no significant differences across eleven categories (**Fig. S6**). The most frequent nASE categories, “partial intron retention” and “annotated intron reduced usage”, were not significantly different. The only exception was the “complex” category, which was observed in four BD cases and was absent in unaffected relatives (*P*=0.04). Notably, several nASE types with potentially higher functional impact, including “annotated intron increased usage”, “complex”, “exon truncation”, and “exon truncation and elongation”, were detected exclusively in BD cases, although their limited numbers warrant cautious interpretation.

**Figure 6:**
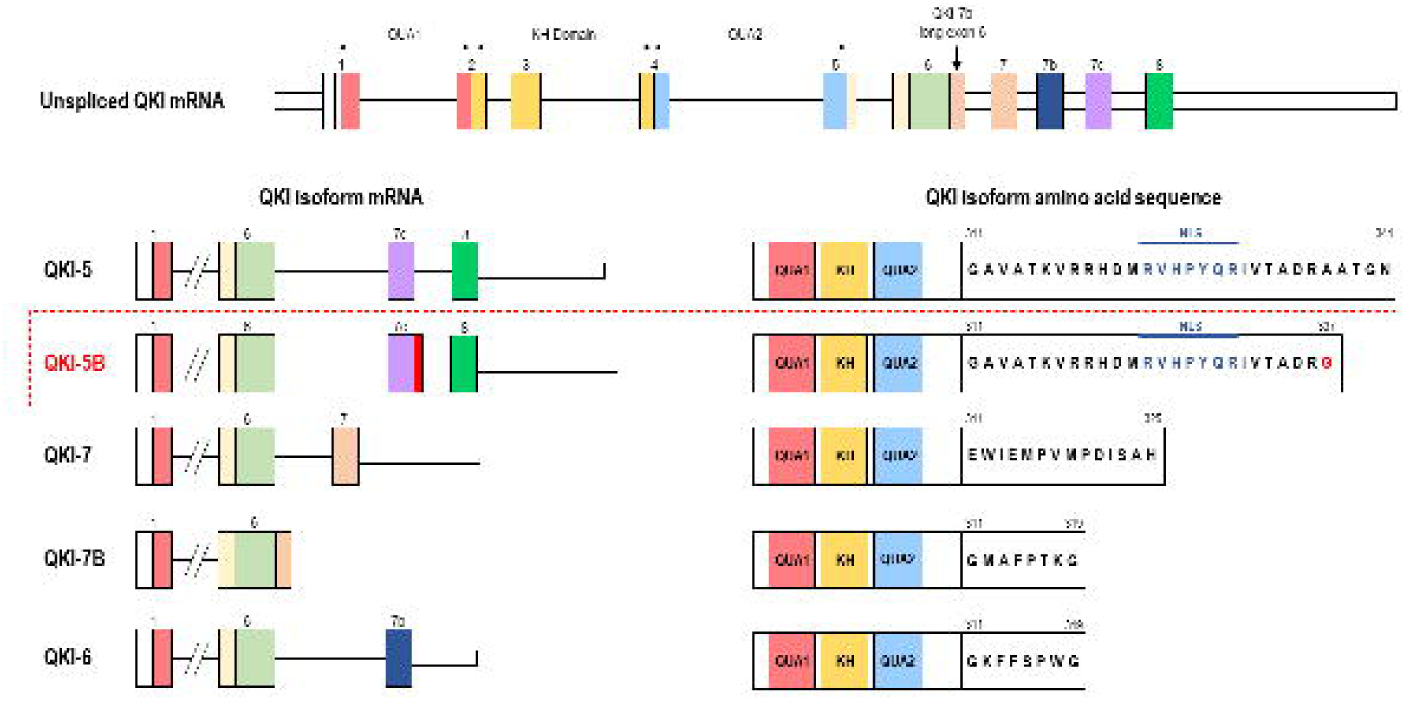
Volcano plots showing nASEs for each affected individual in the DS (family-based). For each sample, the x-axis represents the ΔIntron Jaccard Index across all splice sites, and the y-axis represents the –log10(*P*-value). Red dots indicate identified nASEs with a *P*adj < 0.05.

## DISCUSSION

Herein, we present a comprehensive transcriptome-wide study of alternative splicing in BD. To our knowledge, this is the first study that integrate multiple computational frameworks, use both familial and independent case-control replication cohorts, apply gene and event-level prioritisation approaches, and suggest biological pathways relevant for BD. By applying three distinct TWASA methodologies, alongside novel splicing outlier detection, we captured both predicted group-level splicing dysregulation and novel rare, potentially high-impact aberrant events, providing a multi-layered view of splicing complexity in adults impacted by a psychiatric condition.

We first applied three widely TWASA tools: VT, RM, and MCC. VT maps RNAseq reads to predefined exon–exon and exon–intron junctions, which enables mappability correction and sensitive microexon detection, but does not support the discovery of novel annotated splicing events [52]. RM, similarly to VT, focuses on basic splicing categories, and applies a statistical framework to identify predicted differential splicing events between groups [53]. In contrast, MCC adopts a conceptually distinct approach based on local splicing variations (LSVs), enabling the detection of both basic and complex events, such as tandem cassette exons, orphan junctions, exitrons, and multi-exon spanning events, while also incorporating correction for known and latent confounders [54,55]. Our results highlight substantial heterogeneity across existing TWASA tools, with no single gene or ASE consistently identified across all three approaches. This limited concordance, which has been reported previously [43], likely reflects substantial divergence among splicing detection algorithms and highlights the sensitivity of TWASA results to methodological choices, including differences in splicing unit definitions, estimation of inclusion levels (PSI), statistical models, covariate corrections, and the ability to capture higher order or complex ASE. Rather than reflecting biological inconsistency, this heterogeneity highlights the need for complementary integrative, multi-tool pipelines to capture splicing complexity more comprehensively in psychiatry. While VT and RM identified numerous basic ASEs, only complex ASEs detected by the MCC pipeline showed robust genetic enrichment for BD, but also with SUD and SCZ, confirming the known shared genetic architecture between BD and these two phenotypes [6,67]. Functional enrichment and gene-set association analyses on the subset of genes identified by MCC converge on neurobiologically and behaviourally relevant pathways shared between BD and schizophrenia, such as aggressive behaviour, synaptic function, or abnormal social behaviours typically associated with mood and psychotic disorders [68–72].

Overall, the TWASA analyses suggest that complex splicing alterations, rather than isolated exon inclusion or skipping events, may be more biologically and genetically relevant to psychiatric phenotypes. On this basis, the more recently developed MCC tool was selected as the primary framework for downstream analyses.

After stringent filtering and systematic prioritization, 441 genes identified by MCC had convergent support of gene-level genetic association for BD and putative disease-relevant splicing alterations.

Notably, *DOCK2* emerged as a top candidate, ranked among the strongest candidates based on gene-level association analysis. This gene harbours a genome-wide significant BD GWAS signal, and has been recently identified as a likely causal gene in the PGC4 BD GWAS [6]. *DOCK2* encodes a guanine nucleotide exchange factor (GEF), a class of proteins with established roles in immune signalling, microglial function, and neuronal development, processes increasingly implicated in the aetiology of mood disorders [73–79].

The splicing architecture of *DOCK2* revealed two alternative splicing sub-events: i) a double exon skipping event affecting two non-canonical exons, which was more frequent in individuals with BD and corresponds to the canonical isoform; ii) and an intron retention event occurring more often in unaffected individuals that aligns with a non-annotated isoform leading to a premature stop codon.

This finding represents an example in which subtle shifts in the relative abundance of specific transcripts, particularly those that are less frequent than the main isoforms, may have important biological consequences. Indeed, this may occur in the absence of significant differences in overall gene expression between cases and unaffected individuals. Integrating complex ASE detection with genetic association data can highlight informative molecular mechanism contributing to psychiatry [40].

Prioritized gene subset identified through MCC pipeline underwent replication in an independent case/control cohort, resulting in 98 replicated genes, although modest splicing sub-event level replication was found with only five ASEs. Of those, four ASEs, in *ANKRD36, RBM26, TATDN2,* and *QKI*, showed the same dPSI direction in both datasets. These four genes represent good candidates for BD, all with some degree of genetic association to the disorder.

*ANKRD36* is characterized by an ankyrin domain and modulates the expression of the epithelial sodium channel [80]. A single-nucleus transcriptome-wide association study of the dorsolateral prefrontal cortex in 424 subjects identified *ANKRD36* as candidate gene for depression [81]. *RBM26* encodes a protein involved in RNA processing and nuclear RNA decay, in which recently has been found one of the three significant loci in a Tourette/tic-disorder GWAS meta-analysis, suggesting potential relevance in psychiatry [82]. *TATDN2* is a RNA 3’ exonuclease and endonuclease involved in the degradation of the annealed single-stranded RNA and resolving R-loops [83]. *TATDN2* was one of 23 genes able to differentiate MDD from controls using gene expression data [84]. Finally, *QKI* is involved in RNA binding stability, alternative splicing, and mRNA precursor processing and transport [85]. This gene plays a central role in the central nervous system, especially in oligodendrocyte differentiation and myelinogenesis [86–88], and has previously been implicated in schizophrenia [89].

RNA-binding proteins (RBPs) such as QKI are master regulators of hundreds of transcripts in the brain, and their dysfunction can lead to severe phenotypes, as seen for fragile X syndrome via *FMR1* [90]. RBPs are essential for synaptic plasticity, neurogenesis, and neurodevelopment, and their dysregulation is linked to psychiatric disorders [91,92]. Perturbation of individual RBPs can affect multiple target genes, exerting large effects in downstream regulatory networks. *QKI* has been implicated in schizophrenia by different independent reports [89,93], and in major depression in patients who died by suicide [94]. *QKI* is characterized by multiple alternative splicing isoforms, including QKI-5, QKI-6, QKI-7, and QKI-7B, generated through differential splicing across the exon 6 to 8. These isoforms are expressed in the human frontal cortex, and reduced expression of the QKI-7 and QKI-7B isoforms have been reported in schizophrenia [95]. The QKI-7 isoform, in turn, modulates the expression of downstream oligodendrocyte-related genes [89]. Haloperidol, a typical antipsychotic drug, seems to modulate the QKI-7 isoform, increasing its expression in a human astrocytoma cell line [96]. These findings support the notion that the balance between QKI splice variants, rather than overall QKI expression, plays a critical role in regulating oligodendrocyte and myelin-associated gene networks.

Interestingly, the isoform identified in our study (QKI-5B) retains most of intron 7 and is predicted to encode a 337-amino acid protein, rather than the canonical 341-amino acid QKI-5 isoform. Notably, this variant lacks predicted RSK kinases phosphorylation sites, and RSK proteins are known to play a crucial role in neuronal development [97]. Interestingly, the QKI-5B found in our study is expressed in the brain, and similarly to QKI-5 are the only isoforms with a signal peptide for nuclear localization. In our study, we found higher proportion of this alternative QKI-5B isoform in controls compared to BD cases. We can speculate that this alternative protein isoform may be needed to bind different subsets of transcripts or have other downstream processes at nuclear level, which in BD cases is found less abundant. This evidence supports a likely role for *QKI* in psychotic disorders, whereby alterations in its splicing or expression can exert downstream effects on genes involved in oligodendrocyte maturation and myelination. Taken together, our findings suggest that isoform-specific alterations resulting from alternative splicing may contribute to psychiatric disease mechanisms rather than changes in overall gene expression.

Finally, we did not observe substantial differences in individual categories of nASEs between affected and unaffected individuals within families, although we detected a modest enrichment of “complex” nASEs in cases and a trend towards higher level of “annotated intron reduced usage” in unaffected individuals. Interestingly, a previous work reported that mood stabilizers reduced global intron-retention rates [41], suggesting that intron-related alternative splicing may be related to disease state or treatment.

We did not detect evidence of familial segregation of nASEs, indicating that these events are likely to be driven by individual-specific regulatory variation rather than inherited mechanisms. A small number of nASEs predicted to result in frameshifts and protein truncations were identified both in affected and unaffected subjects. However, the limited sample size precludes definitive conclusions in regard to their pathogenic role in the disease. Our findings suggest that nASE do not play a major role in familial BD. Nevertheless, larger studies will be required to fully assess their potential contribution to BD risk, in a similar manner to studies performed recently for *de novo* DNA variants [10,98,99].

Despite the insights provided by this study, some limitations should be acknowledged: Firstly, while the RNA employed in the familial and case-control cohorts were both blood-derived, cellular heterogeneity of source material specific to each cohort (i.e. LCLs and whole blood, respectively) may have impacted cross-cohort reproducibility. While these tissues offer practical advantages in terms of accessibility, studies such as this are otherwise limited by available brain tissue, thus they may not fully recapitulate splicing patterns seen in the brain [100]. Secondly, the relatively limited sample size in the familial cohort may have reduced statistical power to detect ASE, particularly those of subtle or modest effect size. Thirdly, despite continuing advances in method development, ASE detection remains computationally noisy, and tool-specific biases likely impact set of events identified. Fourthly, despite stringent statistical and coverage filters, isoforms with potential disease relevance may have been missed due to low abundance. Finally, our replication strategy was restricted to genes with prior evidence of genetic association, potentially missing ASEs in novel genes which await discovery.

In summary, this study presents the first comprehensive study of alternative splicing in BD, employing a suite of analytical approaches, GWAS-guided prioritization, replication in an independent cohort, and exploration of novel ASEs. Alteration in the abundance of complex alternative splicing events appear to contribute to the molecular pathology of BD. We identified specific isoforms in candidate genes such as *DOCK2*, *RBM26*, *QKI*, *ANKRD36*, and *TATDN2*. We also highlight the contribution of novel transcriptomic splicing events in BD.

Overall, our findings support the involvement of alternative splicing dysregulation as a potential molecular mechanism contributing to BD. Further studies are required to better characterize this largely unexplored regulatory layer in psychiatric disorders.

## Data Availability

Individual-level data are not available for data sharing. Sharing of summary data (raw count matrix) will be considered upon request.

RNAseq individual-level data are not publicly available for data sharing. Sharing of summary data will be made available upon request. All GWAS summary statistics employed in this work are publicly available at https://pgc.unc.edu/.

## ACKNOWLEDGMENTS

This study was supported by grants: RyC2018-024106-I, PID2020-114996RB-I00, CNS2022-135318, PID2023-149154OB-I00 funded by MICIU/AEI/10.13039/501100011033, FEDER UE, European Union

NextGenerationEU/PRTR and ESF Investing in your future (Toma); the NAB Foundation “Stronger Minds” (Cooper and Fullerton); The Australian National Health and Medical Research Council (NHMRC) Program Grant 1037196 (Mitchell and Schofield); NHMRC Project Grant 1063960 (Fullerton and Schofield); NHMRC Investigator Grants 1176716 (Schofield) and 1177991 (Mitchell); NHMRC & Medical Research Futures Fund Grant 1200428 (Fullerton). García-Ortiz was supported by the Fundación Tatiana Pérez Guzmán el Bueno fellowship, and Dr Romero-Miguel by the Juan de la Cierva fellowship (grant JDC2023-052237-I) funded by MCIN/AEI/10.13039/501100011033 and ESF+. Dr Fullerton was a recipient of the Janette Mary O’Neil Research fellowship and was supported by philanthropic donations from Betty C. Lynch OAM. The funders had no role in the design collection, management, analysis, and interpretation of the data.

## CONTRIBUTION

MMJ and CT conceptualized and designed the study. Statistical and computational analyses were performed by MMJ, IGO. Study supervision was carried out by CT. Funding was obtained by AAC, JMM, PRS, PBM, EBG, and CT. MMJ and CT contributed substantially to drafting the manuscript, and MMJ and DRM prepared the figures. All authors contributed to critical review of the final manuscript.

## COMPETING INTERESTS

JMF received honoraria from Illumina for contribution to a Speakers Bureau in 2023 and had travel expenses paid by Novo Nordisk Fonden in 2023 (not related to this work). No other disclosures were reported. EBG has been a consultant to or has received honoraria or grants from Janssen Cilag, Lundbeck, Otsuka, Pziffer, Servier, Deprexis and Sanoffi. EBG is founder of eB2, and designed MEmind. No other disclosures were reported from the other authors.

## Notes

### Author Declarations

All procedures were conducted in accordance with protocols approved by the University of New South Wales Human Research Ethics Committee (initial approval HREC04144; extensions HREC10078, HC15503, HC16347). For the MadManic cohort ethical approval for handling clinical, digital, and for all aspects of the study was obtained by the CSIC Research Ethics Committee (13/2021; 109/2023; 030/2025) and Fundacion Jimenez Diaz Hospital on behalf of all participating hospitals in this study as part of the Quironsalud Group (15/21; 11/23).

