## Supplementary Material and Figures for "Transcriptome-Wide Alternative Splicing Analysis Implicates Complex Events in Bipolar Disorder"

### Contents


| | |
|---|---|
| Cohorts and clinical assessments | 2 |
| RNAseq sequencing and preprocessing | 3 |
| TWASA computational tools | 4 |
| VAST-TOOLS | 4 |
| rMATS | 6 |
| MAJIQ | 7 |
| MOCASSIN | 9 |
| FRASER | 11 |
| Supplementary Figures | 13 |
| Supplemental References | 20 |

**Cohorts and clinical assessments**

The BD family cohort in this study was selected from a larger cohort of 65 multiplex families recruited at the Mood Disorders Unit and Black Dog Institute at the Prince of Wales Hospital, and the School of Psychiatry, University of New South Wales, Sydney, Australia. Medium-to-large multiplex and extended multigenerational pedigrees were ascertained after initial consultation with a proband with bipolar disorder type I (BD-I). Peripheral blood samples were collected from each pedigree member for DNA extraction and the generation of cell lines by standard laboratory methods. Clinical information obtained from the FIGS, DIGS, and medical records was used to derive best-estimate Research Diagnostic Criteria (RDC) diagnoses under the DSM-IV for bipolar disorder type I (BD-I), bipolar disorder type II (BD-II), schizoaffective disorder-manic type (SZMA), or recurrent unipolar depression (RUD) [1]. All participants provided written informed consent. All procedures were conducted in accordance with protocols approved by the University of New South Wales Human Research Ethics Committee (initial approval HREC04144; extensions HREC10078, HC15503, HC16347).

The Spanish case-control cohort was recruited as part of the Madrid Manic Group (MadManic) cohort, which includes BD-I, BD-II, and SZMA patients clinically assessed at following hospitals and clinical units in the metropolitan area of Madrid (Spain): Hospital Fundación Jiménez Díaz, Hospital General de Villalba; Hospital Rey Juan Carlos, and Hospital Infanta Elena de Valdemoro. Clinical assessments were conducted via face-to-face interviews and diagnoses established accordingly to DSM-V. Clinical data and scales were recorded electronically. Control participants were recruited through two occupational medicine services of the Spanish National Research Council (CSIC) at Autonomous University of Madrid (Cantoblanco Campus) and the CISC national centre in Madrid. Participants provided peripheral blood samples for extractions of DNA, RNA, and plasma. Fresh blood samples were processed within 2–6h and all biospecimens were centralized at Centro de Biología Molecular Severo Ochoa (CBMSO). Ethical approval for handling clinical, digital, and for all aspects of the study was obtained by the CSIC Research Ethics Committee (13/2021; 109/2023; 030/2025) and Fundación Jimenez Díaz Hospital on behalf of all participating hospitals in this study as part of the Quironsalud Group (15/21; 11/23).

**RNAseq and preprocessing**

For the DS, libraries were prepared with Illumina Tru-seq stranded mRNA library kit following the manufacturer's protocol. Prepared libraries were amplified for 13 cycles using the KAPA HiFi HotStart Library Amplification Kit (Roche), and subsequently sequenced on the Hiseq 2500 (Illumina) at The Kinghorn Centre for Clinical Genomics (Garvan Institute of Medical Research, Sydney, Australia) using paired-end sequencing v4.0 chemistry over 125 cycles.

For the RS sample, RNA quality was initially assessed, with a minimum RNA integrity number (RIN) of 5.5 required. Globin depletion was performed with Globin-Zero Gold rRNA Removal Kit. Library was sequenced on NovaSeq X Plus Series (PE150) using the Illumina platform at Novogene GmbH (Novogene, Munich, Germany), with each sample sequenced to a depth of approximately 100 million paired-end reads, generating ~15 Gb of raw data per individual. This high-resolution transcriptomic profiling enables quantification of gene expression, isoform diversity, and the detection of low-abundance transcripts, supporting in-depth analysis of peripheral blood transcriptomes. Raw reads were trimmed using Trimmomatic with the following parameters: SLIDINGWINDOW:4:20 and MINLEN:50. Alignment was performed with Hisat2 using the following parameters: *-q –rna-strandness RF –k 1*.

**TWASA computational tools**

The TWASA was performed through three different computational tools: VAST-TOOLS v2.5.1 (VT), rMATS v4.1.2 (RM), and MAJIQ and MOCCASIN v2.4 (MCC).

**VAST-TOOLS**

VT was executed sequentially using the *align*, *combine*, and *tidy* modules. The *align* module processes RNAseq data from the FASTQ/FASTA files to generate seven types of intermediate files, which serve as input files for the subsequent *combine* module. The *combine* module produces the main ASE output table, which is then simplified by the *tidy* module to retain only the VastIDs and the PSI values for samples with sufficient coverage. All modules were run with default parameters, using the *-sp Hs2* option for the hg38 genome assembly. The transcriptome-wide comparisons of ASE between BD and unaffected relatives were performed using the Wilcoxon test (wilcox.test in R). VAST-TOOLS (VT) is capable of identifying and quantifying four major types of alternative splicing events (ASE): (a) exon skipping (including microexons) (ES), (b) intron retention (IR), (c) alternative donor choices (ALTD), and (d) alternative acceptor choices (ALTA). This software differs from other AS-quantification tools in two main aspects. First, VT is independent from splice mappers and performs a mappability correction by directly mapping RNAseq reads to sets of predefined exon-exon and exon-intron junctions (EEJs). Second, VT relies on its own annotation database (*VASTDB*) that provides splicing quantification for a predefined and fixed set of ASE for each species. This allows VT to assign a unique identifier (VastID) to each ASE and to quantify events consistently across any processed RNA-seq sample. However, this approach prevents the detection of novel ASE. VT is organized into modules that perform different steps in the ASE analysis pipeline: (i) quantify ASE inclusion levels (*align*), (ii) combine these levels into individual tables (*combine* and *tidy*), (iii) perform differential splicing analyses (*compare* and *diff*), and (iv) plot the inclusion levels of ASE of interest (*plot*). Nevertheless, this toolset is not recommended for large datasets. In addition, when the dataset is not large, but it includes five or more replicates, it is recommended to perform standard statistical tests using *vast-tools tidy* output instead of performing the differential splicing analyses with *compare* and *diff* modules. Since this study uses more than five replicates, the modules (iii) and (iv) were not used and are therefore will not be addressed on the following explanation:

(i) vast-tools align

It processes the RNAseq data from FASTQ/FASTA files to generate files with read count information. Reads are mapped to a predefined set of EEJs through different submodules (splice site-based, transcript-based, microexon, and intron retention). For this, it trims the reads into 50 nucleotide (nt) fragments using a 25-nt sliding window. Afterwards, the fragments are aligned using bowtie and only uniquely mapping fragments are considered for the splicing quantification. In addition, *vast-tools align* includes a dedicated submodule for the quantification of microexons of length ≤ 15 nt.

(ii) vast-tools combine and vast-tools tidy

*Vast-tools combine* processes the read files and builds the main VT output table, which includes: (a) the inclusion level, using the Percent Spliced-In (PSI) metric, and (b) a comma-separated array of five scores representing the reliability of the PSI measure, along with normalized read counts supporting inclusion and exclusion.

*Vast-tools tidy* simplifies the output generated by *vast-tools combine*, producing a table that contains only the VastID and the PSI values for samples with a minimum coverage support. For samples not meeting the minimum coverage threshold, PSI values are set to NA.

(iii) Wilcoxon test

After obtaining the table from *tidy*, a Wilcoxon test can be performed to calculate the p-values for deltaPSI values obtained from the ASE analysis, instead of using the *diff* or *compare* modules of VT.

**rMATS**

rMATS uses a hierarchical framework to model exon inclusion levels, denoted as $\psi$ or percentage spliced in (PSI), that accounts for both estimation uncertainty within individual replicates and variability among replicates. It uses a likelihood-ratio test to calculate the P value assessing whether the difference in mean inclusion levels between two sample groups exceeds a user-defined threshold (e.g., $|\Delta\psi| = |\psi_{i1} - \psi_{i2}| > 5\%$ ). When estimating $\psi$, rMATS accounts for uncertainty of each individual sample due to sequencing coverage, and biological or technical variability among replicates. The uncertainty of $\psi$, influenced by the total read count, is modelled using a binomial distribution. For example, in case of a skipped exon, the exon inclusion level can be estimated by using the number of reads supporting the exon inclusion ($I$) and the number of reads supporting the exon skipping ($S$), along with the effective lengths of the inclusion ($l_I$) and skipping ($l_S$) isoforms. Therefore, the exon inclusion level $\psi$ can be estimated as $\hat{\psi} = (I/l_I)/(I/l_I + S/l_S)$. Assuming that the inclusion read count $I$ follows a binomial distribution, with the total read count $n = I + S$, we have

$$I|\psi \sim \text{Binomial}(n = I + S, p = f(\psi) = \frac{l_I\psi}{l_I\psi + l_S(1-\psi)}),$$

where the binomial distribution models the estimation uncertainty of $\psi$ as influenced by the total read count $n$, while the proportion of reads from the exon inclusion isoform is represented by the length normalization function $f(\psi)$ that normalizes the exon inclusion level by the effective lengths of the isoforms.

Within a sample group, the exon inclusion levels vary among replicates. This variability is modelled by random effects in a mixed model. For two sample groups $j = 1,\ 2$, with $M_1$ $(k = 1,\ \ldots\ , M_1)$ and $M_2$ $(k = 1,\ \ldots\ , M_2)$ replicates respectively, for each exon $i$, rMATS estimates the group mean of exon inclusion levels of groups 1 and 2 ($\psi_{i1}$ and $\psi_{i2}$) as fixed effects. Then, it assumes that the logit transformation of exon inclusion levels in individual replicate $k$ ($\psi_{ijk}$) follows a normal distribution with the logit of the group mean ($\psi_{ij}$) and the group variance ($\sigma_{ij}$) for modelling the variability among replicates:

$$\text{logit}(\psi_{ijk}) \sim \text{Normal}(\mu = \text{logit}(\psi_{ij}), \sigma^2 = \sigma_{ij}^2).$$

In sum, rMATS jointly models estimation uncertainty and inter-replicate variability using a hierarchical model. Differential splicing is assessed using a likelihood-ratio test to evaluate whether the difference of the group mean between the two sample groups exceeds a user-defined threshold $c$, against the null hypothesis $|\Delta\psi_i| = |\psi_{i1} - \psi_{i2}| \leq c$.

**MAJIQ**

MAJIQ quantifies AS using percent spliced in (PSI, denoted by $\psi$), defined as the relative ratio of isoforms including a specific splicing junction or retained intron. Unlike tools that focus only on canonical ASE (e.g., exon skipping or intron retention), MAJIQ can quantify more complex splicing patterns by using local splicing variations (LSVs) as the unit of analysis. The developers define LSVs as “splits (multiple edges) in a gene splicegraph where several edges either come into or from a single exon, termed the reference exon”, which allow capturing events that involve more than two alternative junctions, avoiding the restriction of modelling ASE as strictly binary decisions of the spliceosome.

The MAJIQ workflow begins with MAJIQ builder, which constructs a splicegraph for each gene combining transcript annotations with aligned RNAseq reads, including unannotated elements. Optionally, a read correction step can be performed using MOCCASIN, which is explained further below. Afterwards, splicing quantification is performed using MAJIQ quantifier, which supports three analysis modes: (i) *psi*, (ii) *deltapsi*, and (iii) *heterogen*. Depending on the selected mode, each LSV edge, corresponding to a splice junction or intron retention, is quantified either in terms of its relative inclusion (PSI, $\psi \in [0,1]$) or by changes in its relative inclusion between two conditions (dPSI, $\Delta\psi \in [-1,1]$ ). MAJIQ employs a Bayesian model that accounts for the number of reads and other factors (such as read distribution across genomic locations and read stacks) to compute the posterior distributions over the (unknown) inclusion level ($\mathbb{P}(\psi)$) when using *psi* mode, or the changes in inclusion levels between conditions ($\mathbb{P}(\Delta\psi)$) when using *deltapsi* mode. This Bayesian framework allows the computation of confidence measures, such as the probability that the inclusion change $|\Delta\psi|$ exceeds a threshold C ($\mathbb{P}(|\Delta\psi| > C)$), as well as the expectation over the computed posterior distributions ($\mathbb{E}[\psi]$, $\mathbb{E}[\Delta\psi]$ ). This leads to the *heterogen* mode, that is the result of the implementation of additional test statistics. This mode quantifies PSI for each sample individually and then applies robust rank-based test statistics (TNOM, InfoScore, or Mann-Whitney U/Wilcoxon), rather than assuming a shared (hidden) PSI value across samples.

**MOCCASIN**

MOCCASIN algorithm adjusts read rates to remove confounding variation. To do this, MOCCASIN uses the read rates matrix and the design matrix (a list with the confounding factors and covariates). The read rate input data $R$ is a matrix such that each entry $R_{m,k}$ represents the read rate for a splice junction $m \in [1 \dots M]$ in sample $k \in [1 \dots K]$. The design matrix specifies sample-to-condition assignments and is partitioned into confounders $C$ and variables-of-interest/non-confounders $V$. If the known confounders have additional learned factors of unwanted variation, the column $C$ is divided into known $N$ and unknown $U$ confounders.

MOCCASIN assumes that the junctions in the read table are grouped into LSVs, identifying each junction row $m$ with its matching LSV $S(m) = l \in [1 \dots L]$. For each LSV $l$ and sample $k$, MOCCASIN first computes the total read rates over junctions in the LSV ($T_{l,k} = \Sigma_m R_{m,k} s.t. S(m) = l$). Then, the total read rate per LSV in each sample is scaled to the median total read rates for that LSV across all samples, such that the scaled read rates per LSV maintain the same PSI:

$$\hat{T_l} = median_k T_{l,k}$$

$$(\widehat{R_{m,k}}) = R_{m,k} \Omega_{l,k}$$

Where $\Omega_{l,k} = \frac{\hat{T_l}}{T_{l,k}}$ is the scaling factor. After scaling, a function *F()* over the read rate $(\widehat{R_{m,k}})$ is modelled as a linear combination of covariates with homoscedastic noise:

$$F(\widehat{R_{m,k}}) = \alpha_m + \boldsymbol{N}_k \cdot \gamma_m + \boldsymbol{U}_k \cdot \delta_m + \boldsymbol{V}_k \cdot \eta_m + \epsilon_{mk} \ (1)$$

Where $\alpha_m$ is the intercept for junction $m$; $\gamma, \boldsymbol{\delta}, \boldsymbol{\eta}$ are the coefficients corresponding to N, U, and V, respectively; and $\epsilon_{mk} \sim N(0, \sigma^2)$ is assumed to be homoscedastic gaussian noise over the scaled read rates. MOCCASIN includes two implementations for the function $F$. The simpler default function is a unity transformation $F(x) = x$ which translates to a linear model over the scaled read rates. The second is a smoothed towards zero log (STZL) transformation:

$$F(x) = ln(x)\, s.t. x > 2 \ (2)$$

$$F(x) = ax^2 + bx s.t. 0 \leq x \leq 2$$

$$F^{-1}(x) = e^x s.t. x \geq ln(2)$$

$$F^{-1}(x) = \frac{1}{2a}() - b + \sqrt{(b^2 + 4ax)} s.t. 0 \leq x < ln(2)$$

Where:

$$a = \frac{1}{4}(1 - ln(2))$$

$$b = ln(2) - \frac{1}{2}$$

The STZL transformation allows for a log-linear model where the effect of confounder has a multiplicative effect on read rates. This approach avoids exploding negative values for low read rates, avoiding instability at low read counts, while it does not cause variable deviation from the theoretically desirable log transformation for high read rates.

The simple read rate model allows MOCCASIN to derive estimates of $\hat{\alpha}_m, \hat{\gamma}_m, \hat{\delta}_m, \hat{\eta}_m$ through simple OLS regression which can be easily parallelized across LSVs. The corrected read rates $R^*{}_{m,k}$ are then computed as:

$$R'_{m,k} = F^{-1}(,)\, C(,)\, F(,)\, \widehat{R_{m,k}}(,) - N_k \cdot \gamma_m - U_k \cdot \delta_m(,)\,(,); T'_{l,k} = \Sigma_m R'_{m,k}\, s.t. S(m) = l(,)$$

$$R^*{}_{m,k} = \frac{T_{l,k}}{T'{}_{l,k}} R'{}_{m,k}$$

Where *C(x) = x if x > 0* otherwise *C(x) = 0* is a simple clipping function to avoid too low (negative) read rates after confounders effect removal. The normalization factor $\frac{T_{l,k}}{T'{}_{l,k}}$ is set to recover the same total level of read rates per LSV. The adjusted read $R^{*}_{m,k}$ rates are reported back as the output for any downstream algorithm.

Additionally, in order to learn unknown confounders, denoted by the matrix U, MOCCASIN uses a procedure similar to the one used by RUV.

**FRASER**

FRASER identifies splicing outliers by using and modelling split and unsplit reads detected at all splice sites in the RNA-seq data. Afterwards, it computes pvalues using a beta-binomial model and controls for unknown covariation between samples using a denoising-autoencoder.

Initially, FRASER modelled three metrics from split and unsplit reads: (1) the percent spliced in for splice acceptor site usage ($\psi_5$); (2) the percent spliced in for splice donor site usage ($\psi_3$); and (3) the splicing efficiency ($\theta$). To reduce the number of minor outliers, FRASER 2.0 implemented the Intron Jaccard Index (J), a new splicing metric that also incorporates both split and unsplit reads. This metric is defined as "the proportion of reads supporting the splicing of an intron of interest among all reads associated with either splice site of the intron" and is modelled using the same beta-binomial autoencoder approach. Formally, J represents the Jaccard index between the sets of donor-associated reads $D$ and acceptor-associated reads $A$. For a given sample $i$ and intron $j$, the donor-associated set $D_{ij}$ consists of split reads that share the same donor site as intron $j$, along with all unsplit reads that span the exon-intron boundary at that donor site of sample $i$. Similarly, the acceptor-associated set $A_{ij}$ is defined for the split and unsplit reads located at the acceptor site of intron.

The Intron Jaccard index $J_{ij}$ for sample $i$ and intron $j$ is calculated as follows:

$$J_{ij} = \frac{|D_{ij} \cap A_{ij}|}{|D_{ij} \cup A_{ij}|} = \frac{s_{ij}}{\sum_{d \in L_j} s_{id} + \sum_{a \in R_j} s_{ia} + \sum_{t \in \{d_j, a_j\}} u_{it} - s_{ij}},$$

where $s_{ij}$ corresponds to the count of split reads mapping to intron $j$ in sample $i$, $d_j$ is the donor site of intron $j$, $a_j$ is the acceptor site of intron $j$, $L_j$ is the set of introns using $d_j$, $R_j$ is the set of introns using $a_j$, and $u_{it}$ denotes the count of unsplit reads spanning the exon-intron boundary at a splice site $t$.

After computing J, a denoising autoencoder is employed to model and account for unknown covariation across samples. The autoencoder is provided with an input matrix X, which consists of the logit-transformed splice metrics of each intron $i$ and sample $j$ by using the following formula:

$$x_{ij} = \text{logit}\left(\frac{k_{ij} + \alpha}{n_{ij} + 2 \cdot \alpha}\right)$$

In the expression, $k_{ij}$ and $n_{ij}$ represent the numerator and the denominator of the Intron Jaccard index of intron $j$ in sample $i$. The parameter $\alpha$ corresponds to a pseudocount introduced to prevent division by zero or the logarithm of zero.

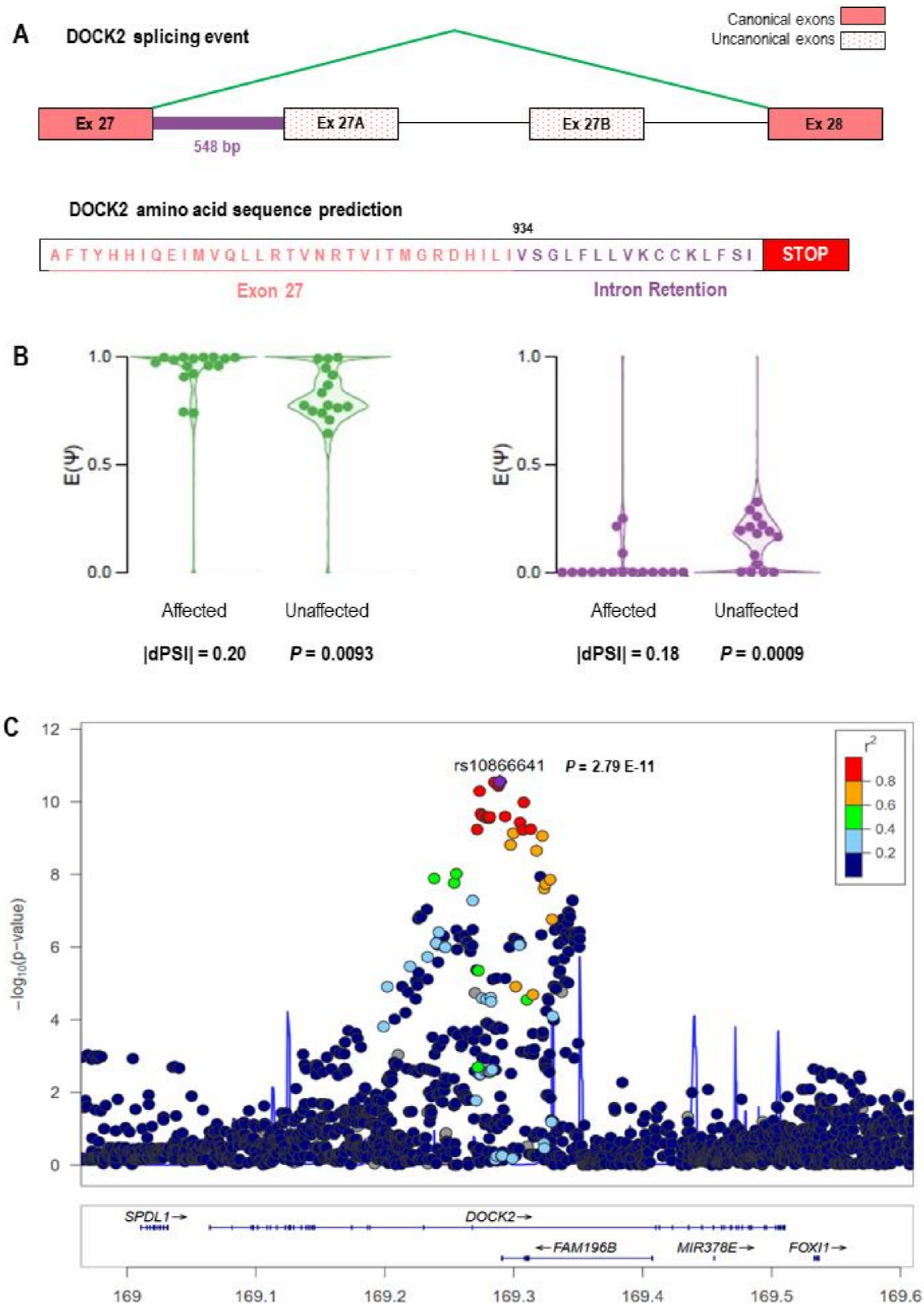


**Supplementary Figure 1**: Representation of ASE identified in the *DOCK2* gene. **A)** Representation of the sub-events identified from exon 27 (hg38/chr5:169,840,757-169,840,852) to exon 28 (hg38/chr5:169,934,652-169,934,871). Exon skipping of exons 27A (hg38:chr5:169,841,401-169,841,535) and 27B (hg38:chr5:169,87,5226-169,875,877) is shown in green, which is more frequent in BD patients; whereas intron retention between exon 27 and exon 27A is shown in purple and is more frequent in unaffected individuals. **B)** Violin plots comparing the distribution of PSI (E(Ψ)) between exon skipping sub-event (green) and intron retention sub-event (purple); each dot represents one individual. Each violin plot represents a significant sub-event of the ASE from exon 27 to exon 28 (WILCOXON P<0.01). **C)** Evidence of genetic association of *DOCK2* region from GWAS for BD (hg19), where each dot represents a SNP plotted by genomic position and –log10(P-value). SNPs are coloured according to linkage disequilibrium ($r^2$) with the leading SNP (rs10866641), and gene annotations across the locus are indicated below.

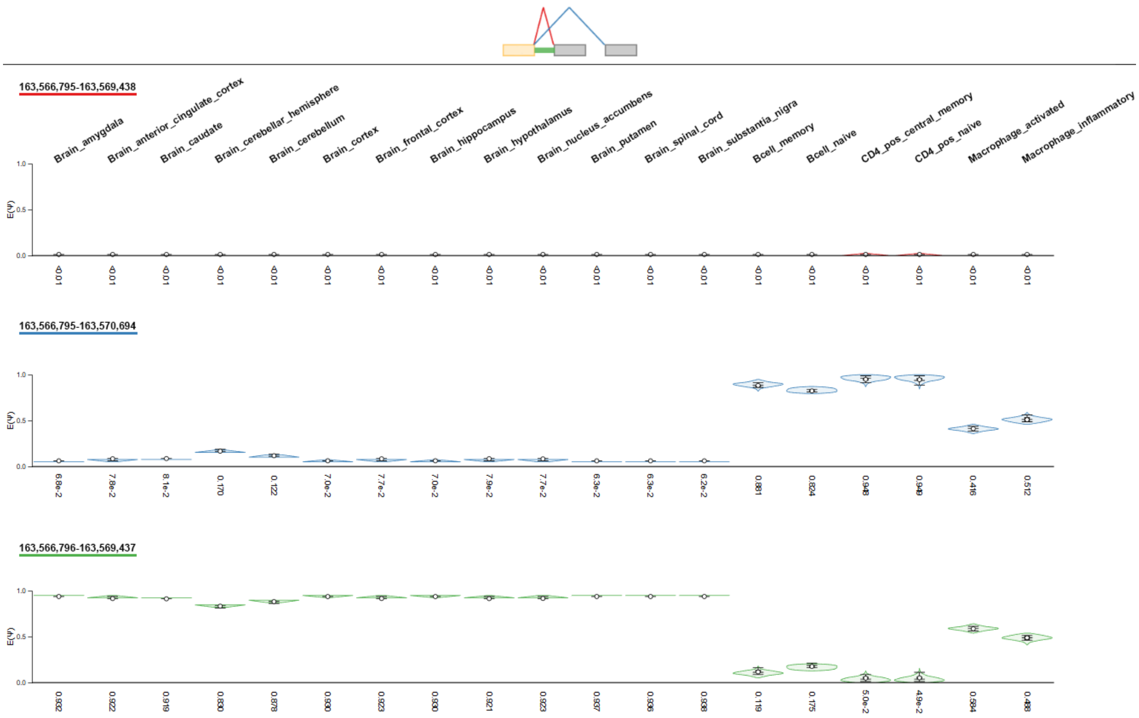


**Supplementary Figure 2**: Violin plots of blood and brain normal tissues from MAJIQlopedia for the event at *QKI* gene (isoform *QKI-5B*; in green) identified and replicated in the independent case-control cohort.

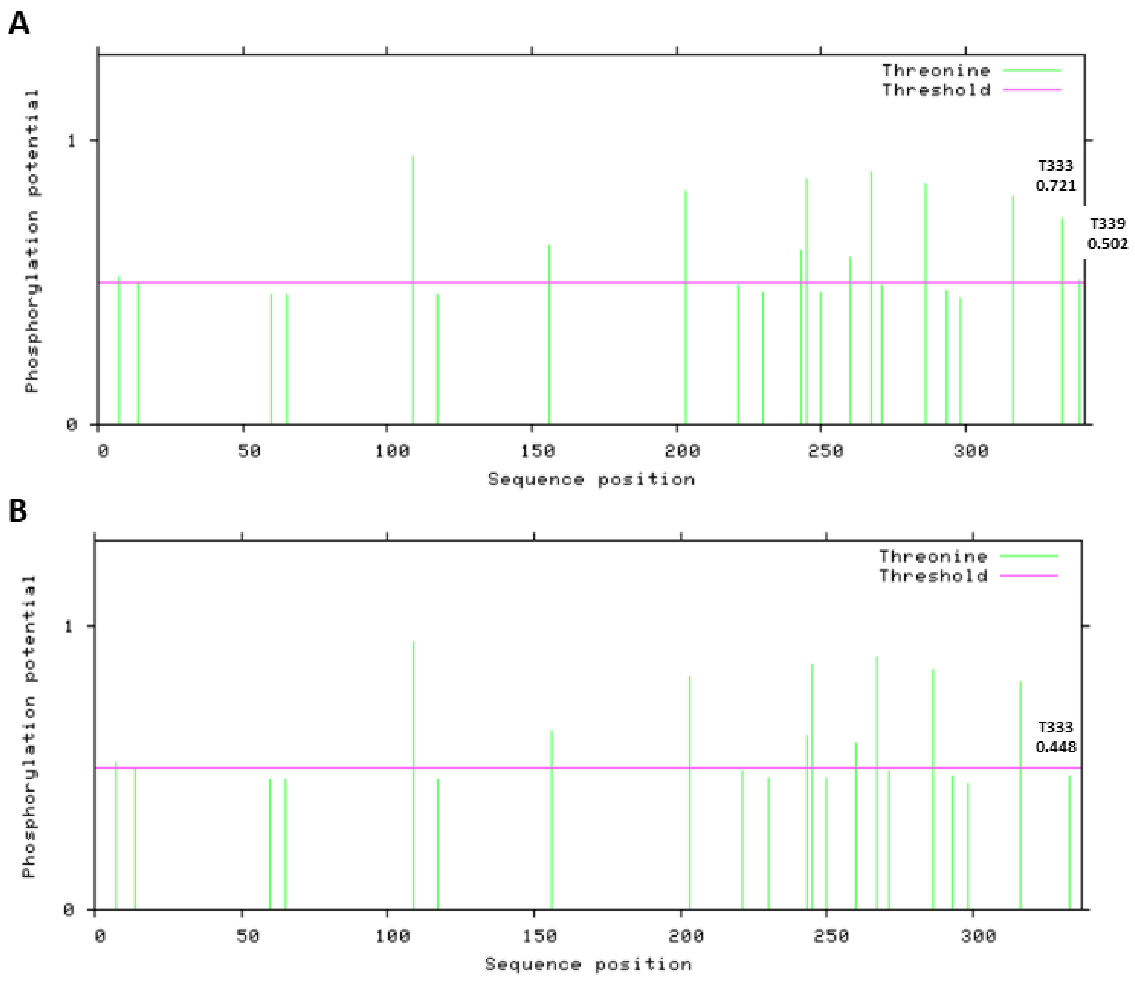


**Supplementary Figure 3**: Spike plots representing the prediction of threonine phosphorylation potential sites across QKI-5 (top) and QKI-5B (bottom) protein sequence. **A)** Predicted phosphorylation sites of canonical isoform QKI-5. **B)** Predicted phosphorylation sites of the alternative isoform described and replicated in this study (QKI-5B). Vertical spikes indicate predicted phosphorylation sites at specific sequence positions, with spike height reflecting the predicted phosphorylation potential. The horizontal line indicates the prediction threshold.

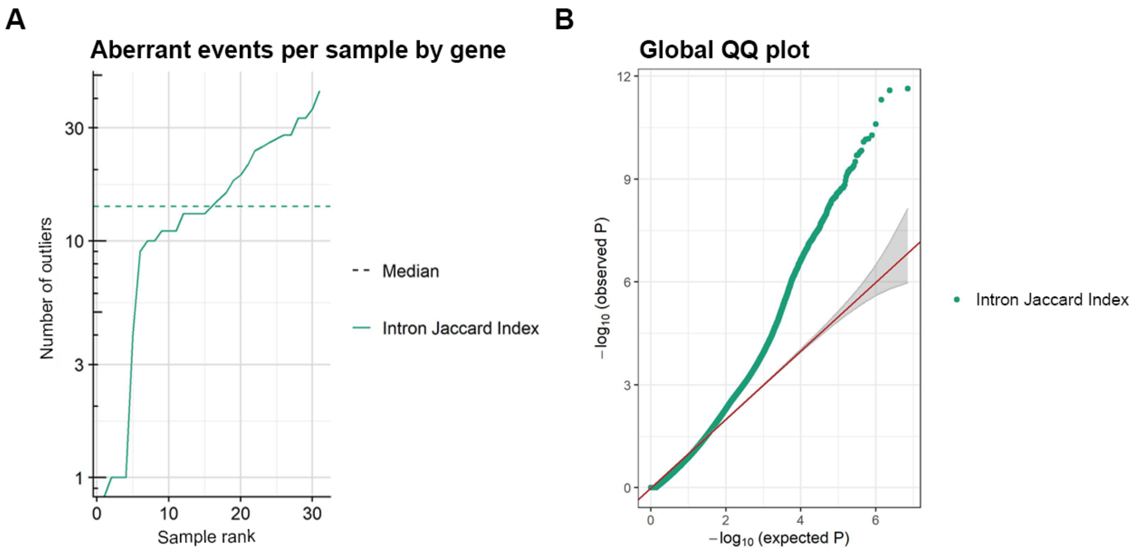


**Supplementary Figure 4**: **A)** Ranked line plot showing the number of nASE per sample, filtered by ΔPSI > 0.05 and Padj < 0.05, ranked from the lowest to the highest number of nASEs. **B)** QQ-plot showing the distribution of significance for nASEs. The x-axis represents -log10(expected pvalue) and the y-axis represents -log10(observed pvalue). Each dot corresponds to a single dnASE.

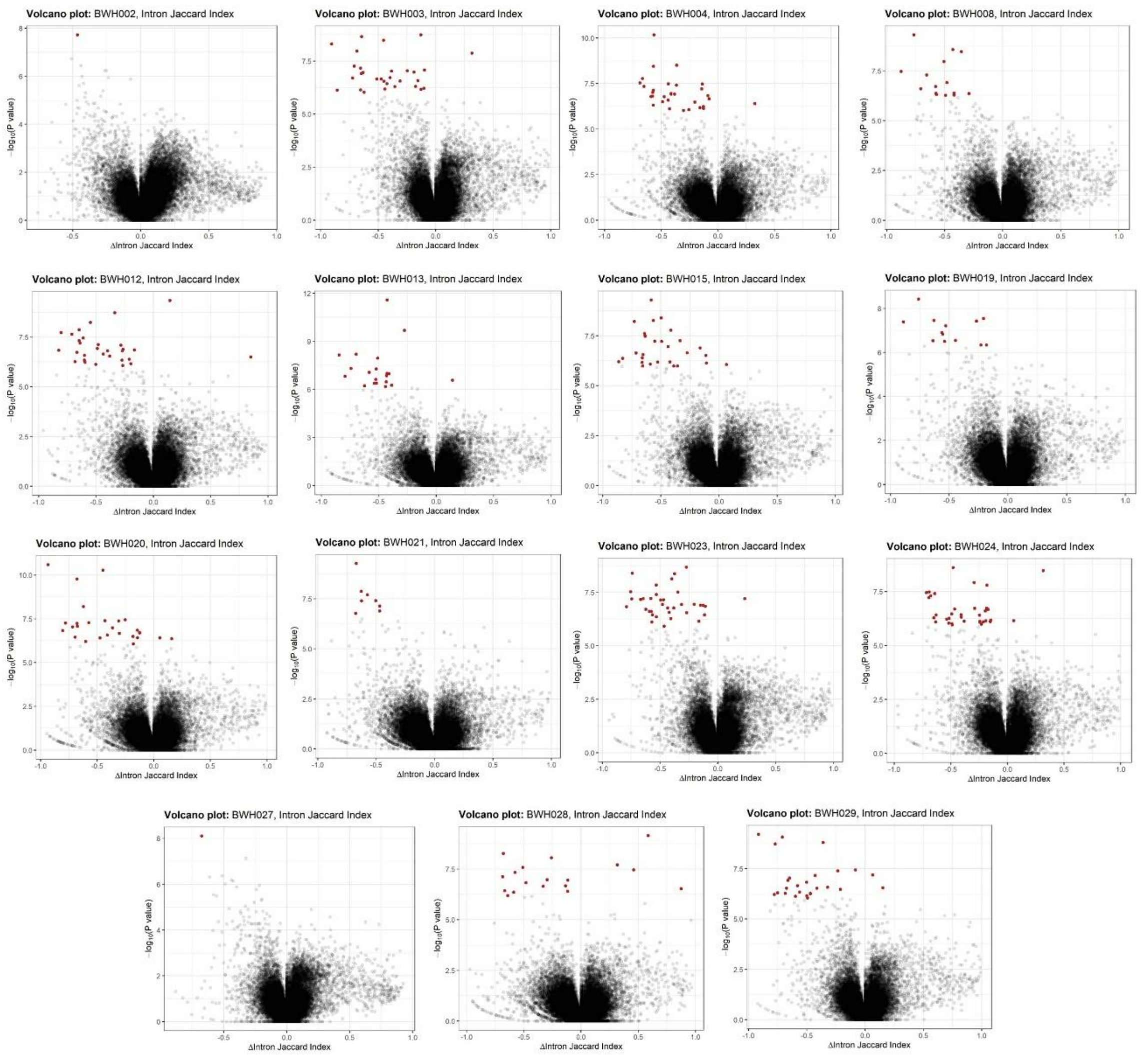


**Supplementary Figure 5**: Volcano plots showing dnASEs for each unaffected individual. For each sample, the x-axis represents the ΔIntron Jaccard Index across all splice sites, and the y-axis represents the –log10(*P*-value). Red dots indicate identified dnASEs with a *P*adj < 0.05.

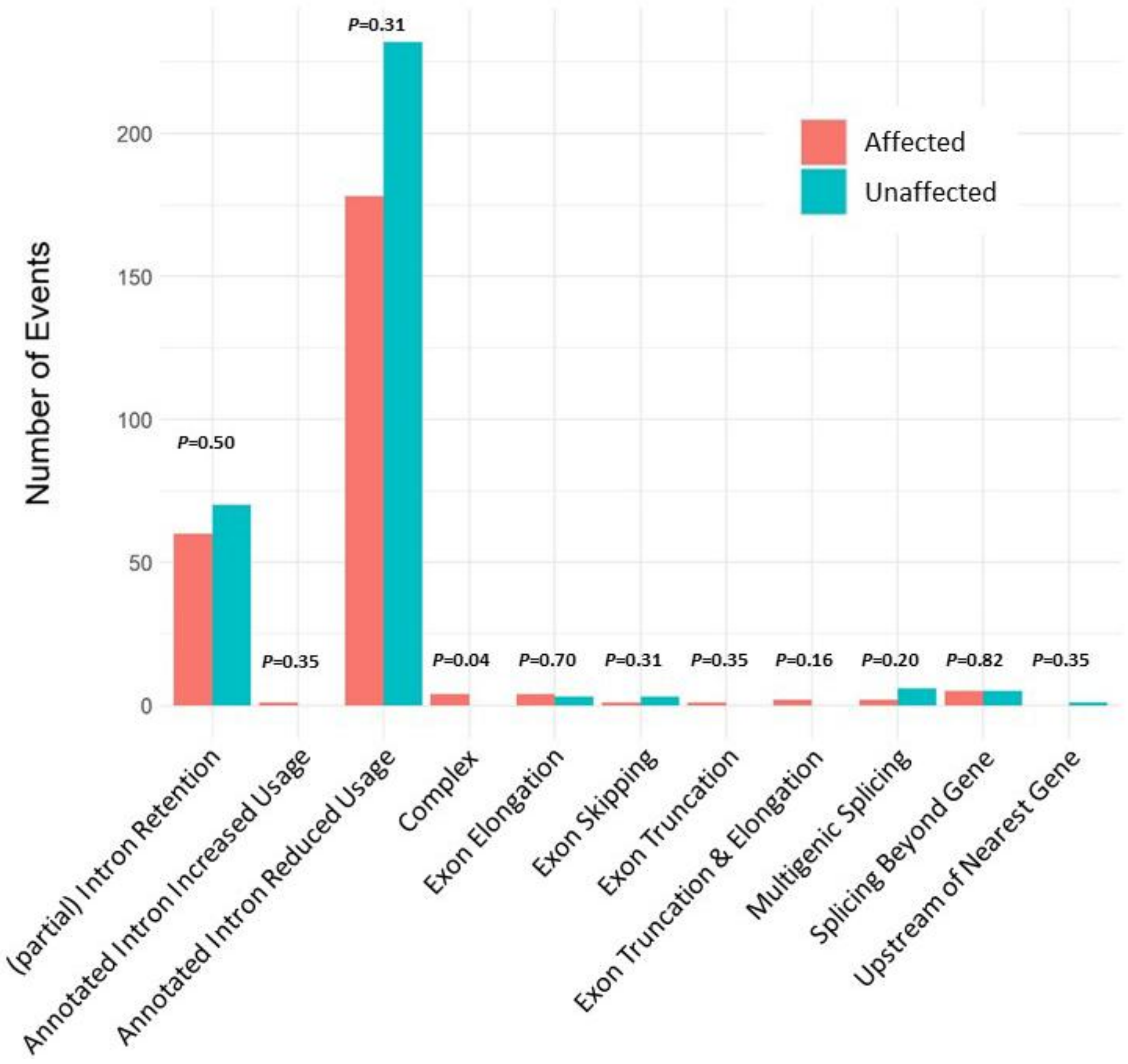


**Supplementary Figure 6**: Bar plot showing the number of dnASEs by event type in affected and unaffected individuals. Bars in coral represent affected individuals, and bars in blue unaffected individuals from the family-based cohort. For each category on the x-axis, the *P-value* from a Wilcoxon test is indicated, assessing differences between the two groups for each event type.
